# Piloting Forensic Tele-Mental Health Evaluations of Asylum Seekers

**DOI:** 10.1101/2020.04.15.20063677

**Authors:** Aliza S. Green, Samuel G. Ruchman, Craig L. Katz, Elizabeth K. Singer

## Abstract

**Problem:** Forensic mental health evaluations can provide critical evidence in the legal cases of individuals seeking asylum and other forms of protected immigration status. While the number of academically affiliated medical human rights programs has increased in recent years, there is still substantial unmet need for pro bono evaluations throughout the United States, especially for individuals in detention.

**Approach:** The Mount Sinai Human Rights Program launched its pilot Remote Evaluation Network in September 2019, with the aim to coordinate forensic mental health evaluations by telephone or video call for individuals who are unable to access in-person services. The authors recruited mental health clinicians from across the country, trained them in best practices in conducting forensic evaluations using telehealth platforms, and coordinated pro bono mental health evaluations of individuals seeking immigration relief. Remote forensic services have been a particularly relevant solution in the context of the COVID-19 pandemic.

**Outcomes:** The Remote Evaluation Network consists of seventeen active evaluators. From December 2019 to April 2020, the pilot program has coordinated fifteen forensic evaluations of individuals seeking asylum and other forms of protected immigration status in six different states. All clinicians participated in a training module on best practices in conducting forensic evaluations by telephone; respondents to optional pre-and-post-module surveys reported an increase in comfort level with conducting telephonic evaluations after participating in the module.

**Next Steps:** We will formally evaluate this pilot program’s services by assessing the quality of medico-legal affidavits from telephonic evaluations, tracking legal outcomes and qualitative feedback from attorneys, and investigating the acceptability of telephonic mental health evaluations among legal professionals. Future directions include expansion to new geographies, including individuals affected by the Migrant Protection Protocols or “Remain in Mexico” program.

## Manuscript

### Problem

Forensic medical and mental health evaluations, and the documentation of their conclusions in medico-legal affidavits, can provide important evidence in asylum seekers’ pursuit of immigration relief in the United States. However, research suggests that there is a substantial unmet need for trained clinicians to conduct pro bono medical and psychological evaluations of individuals seeking protected immigration status.^1^ Non-governmental organizations (NGOs), including Physicians for Human Rights and torture treatment programs that provide longitudinal care, have helped increase access to forensic evaluations significantly throughout the U.S. Academically affiliated medical human rights programs have also proliferated, but the approximately twenty student-run clinics are predominantly located in the Northeast, with a handful in the Southeast and on the West Coast.^2^ Many areas of the country remain underserved, and our program continues to receive forensic evaluation requests— particularly for individuals in immigration detention—from attorneys who are unable to coordinate in-person evaluations.

Recent changes in federal immigration policy suggest that it may become even more difficult for asylum seekers to access in-person forensic services. Immigration detention has increased dramatically in recent years: more than 35,000 individuals are currently in Immigration and Customs Enforcement (ICE) and Customs and Border Protection custody.^3^ Since U.S. Citizenship and Immigration Services’ “last in, first out” policy was enacted in 2018, newly arrived asylum seekers’ claims have been adjudicated in an expedited fashion, sometimes within a matter of weeks.^4^ As a result, attorneys have less time to request and prepare evidence, including forensic evaluations.^5^ In an unpredictable policy environment, flexible solutions are needed to continue expanding access to pro bono forensic evaluations for vulnerable asylum seekers. This manuscript describes our program’s response to these challenges by detailing the development and early outcomes of a tele-mental health pilot. Peer programs may learn from our program’s experiences, especially in the context of the recent shift to telehealth necessitated by the COVID-19 pandemic.

### Approach

In September 2019, the Mount Sinai Human Rights Program (MSHRP) launched the Remote Evaluation Network. This pilot project aims to coordinate pro bono forensic mental health evaluations, by telephone or video call, of individuals seeking asylum or other forms of protected immigration status in the U.S. The remote network is an expansion of MSHRP, a program of the Icahn School of Medicine at Mount Sinai which facilitates forensic evaluations to document the physical and psychological sequelae of human rights abuses and explain these findings to adjudicators of immigration claims. MSHRP coordinates nearly 200 such forensic evaluations of asylum seekers annually in New York City and provides continuity medical care and social services. Three factors prompted the development of the remote network: (a) an increase in requests for evaluations outside of the New York City area, especially for individuals in detention or in areas without human rights programs; (b) anecdotal feedback from attorneys about the efficacy of initial telephonic evaluations; and (c) an MSHRP study which showed that affidavits documenting telephonic mental health evaluations were comparable in quality to affidavits resulting from in-person evaluations.^6^ Encouraged by this early evidence, the pilot program was launched to provide a high-quality alternative for individuals who—due to location, limited local resources, or time constraints in their legal proceedings—are unable to access “gold standard” in-person forensic medical services.

The objectives of this ongoing pilot are to:

1. Recruit a cohort of mental health evaluators from across the U.S. and train them in best practices for conducting forensic evaluations by telephone and video call;
2. Forge partnerships with pro bono legal service providers, particularly in low-resource geographies;
3. Coordinate remote, forensic mental health evaluations for individuals seeking asylum or other forms of immigration relief.

### Recruitment and Onboarding

Clinicians were recruited through peer referrals and direct outreach to NGOs, legal services organizations, and listservs for immigrant and refugee healthcare providers. We first recruited evaluators with significant experience conducting mental health evaluations of asylum seekers. To ensure quality and standardization of the pilot’s work product, experienced evaluators submitted redacted sample affidavits for review by MSHRP faculty leadership.

We also aimed to harness the enthusiasm of clinician-advocates without forensic experience, many of whom were galvanized by the rapidly changing landscape of national immigration policy. All prospective evaluators attended approved general trainings on forensic evaluation of asylum seekers. New evaluators were also required to shadow experienced mentor evaluators and prepare affidavits under supervision before conducting evaluations independently.

All evaluators, regardless of prior experience, were required to participate in our online training module covering best practices for telephonic evaluations (Table 1). The online module was designed by a forensic psychiatrist who has conducted numerous evaluations of asylum seekers, both in person and remotely. The training’s content was structured around questions about conducting telephonic evaluations, solicited from MSHRP’s evaluator network, and incorporated input from a partner legal service organization. The module focused on telephonic rather than video evaluations because many detention centers permit only telephone access. We will continue to revise this module based on participant feedback, with future iterations addressing management of trauma triggers and post-evaluation distress in clients and the navigation of telehealth regulations.

**Table 1:** Summary of the Remote Evaluation Network’s Online Training Module Content

| Topic | Key Points |
| --- | --- |
| **Research Findings** | *Affidavit comparison:* The Mount Sinai Human Rights Program published a study comparing affidavits resulting from in-person and telephonic evaluations, grading them on a rubric based on the *Istanbul Protocol*. There was no difference between telephonic and in-person evaluations for 26 of 30 criteria. Common differences included assessment of general appearance and psychomotor retardation. There was no difference in distribution of Post-Traumatic Stress Disorder or Major Depressive Disorder.<br>*Evaluator experiences:* Although evaluators with telephonic experience noted that lack of visual cues made establishing rapport and assessing mental status more difficult, they expressed comparable ability to diagnose individuals and testify in a client's trial. |
| **Best Practices** | *Before the interview:* Evaluators should consider what preparation is needed, including setting and equipment, electronic surveys emailed in advance, and location of a language interpreter. Sending a photo of the evaluator in advance may help overcome the barrier to rapport building.<br>*At the beginning of the interview:* Evaluators should orient the evaluatee to the purpose, format, and degree of confidentiality of the evaluation. Acknowledge limitations of telephonic modality and ask for assistance of the evaluatee and interpreter, if co-located, in communicating distress, discomfort, and need for breaks. Consider asking the evaluatee to describe their settings, including the level of privacy.<br>*During the interview:* Evaluators should minimize distractions by sitting away from their computer even if using it to take notes. Interludes in language interpretation can be beneficial for taking notes and collecting thoughts.<br>*At the end of the interview:* Evaluators should consider sharing impressions with the evaluatee and providing psychoeducation. Evaluators may also provide recommendations for follow up. |
| **Common Concerns** | *Loss of non-verbal cues:* Evaluators can still rely on audible cues (e.g., crying, long pauses) to guide their interviewing and help assess mental status.<br>*Ability to assess credibility or truthfulness:* Even for in-person evaluations malingering is usually not an issue because individuals have already been vetted by experienced immigration attorneys.<br>*Building a therapeutic alliance:* The goal of the telephonic evaluation is forensic, unfortunately not therapeutic. Informal attorney feedback suggests that individuals often find the experience to be therapeutic despite the barriers.<br>*Communicating via interpreter:* For telephonic evaluation, the communicating by interpreter can actually be advantageous. An interpreter co-located with a client can provide visual data. Pauses for interpretation also make note taking easier.<br>*Confirming client identity:* Similar to in-person evaluations, evaluators should rely on immigration attorneys to confirm the time and place of evaluations. |

### Legal Outreach

We contacted organizations on the Department of Justice’s national list of pro bono legal service providers to assess their clients’ need for forensic evaluations and inquired about their prior experiences with, interest in, and perceived barriers to coordinating remote evaluations.

Our program prioritized working with organizations operating in low-resource geographies, which we defined as regions with lower-than-average asylum grant rates and limited access to forensic medical services. The Remote Evaluation Network was designed to complement existing local resources in areas with unmet need. As we forged partnerships in new regions, we engaged in collaborative discussions with peer programs to ensure our work was not redundant and would not create additional silos. Evaluation requests originated from legal service providers that we initially contacted, pro bono attorneys who learned about our remote services from colleagues, and email listservs of healthcare providers.

### Case Coordination

The remote network uses the case management tools and workflow of MSHRP’s New York-based service-delivery model. Student clinic managers coordinate case logistics and facilitate communication between attorneys and evaluators. For each remote case, considerations include secure telehealth platforms, privacy of evaluation rooms, out-of-state licensure, and client safety. Attorneys are sent an outcome form at regular intervals to collect information on the legal status of the case and qualitative feedback on the role of the evaluation in the asylum process.

## Outcomes

### Capacity Building

From September 2019 to April 2020 seventeen clinicians onboarded with the remote network who were able to accept evaluation requests at the time of this manuscript’s submission. All seventeen individuals had previous experience conducting forensic mental health evaluation of asylum seekers. Eight clinicians with a range of prior forensic medical experiences are waiting to shadow an experienced clinician and prepare an affidavit under supervision in order to complete their onboarding process.

### Remote Evaluations

Since December 2019, the Remote Evaluation Network has completed fifteen evaluations of individuals seeking protected immigration status, with six more cases in the scheduling process. Evaluees were located in six different states and most were seeking asylum (n = 11, 73%) (Table 2). Most individuals were self-identified cis-gender men (n = 11, 73%), originated from Central American or Caribbean countries (n = 13, 87%), and more than half of the clients (n = 8, 53%) were 18-35 years old. Thirteen clients (87%) were adults in ICE detention facilities or other correctional centers. Most evaluations in the network were conducted by telephone (n = 12, 80%). One non-detained minor and two adults received video evaluations.

**Table 2:** Characteristics of Clients Served by the Remote Evaluation Network (n = 15), December 2019 to April 2020

|  | Total (n = 15) | Percent of Total (Rounded) |
| --- | --- | --- |
| **Age (years)** |  |  |
| Minor, < 18 | 1 | 7% |
| Adult, 18-35 | 8 | 53% |
| Adult, >35 | 6 | 40% |
| **Gender** |  |  |
| Trans-gender woman | 1 | 7% |
| Trans-gender man | 0 | 0% |
| Cis-gender woman | 3 | 20% |
| Cis-gender man | 11 | 73% |
| **Region of Origin** |  |  |
| Caribbean | 6 | 40% |
| Central America | 7 | 47% |
| East Asia and Pacific | 1 | 7% |
| Sub-Saharan Africa | 1 | 7% |
| **Primary Type of Protected Status Sought** |  |  |
| Asylum | 11 | 73% |
| Cancellation / Withholding of Removal | 2 | 13% |
| United Nations Convention Against Torture | 1 | 7% |
| T Nonimmigrant Status (T Visa) | 1 | 7% |
| **Current Region in United States** |  |  |
| Midwest | 2 | 13% |
| Northeast | 5 | 33% |
| South | 7 | 47% |
| West | 1 | 7% |
| **Location** |  |  |
| Community | 2 | 13% |
| Detention | 13 | 87% |
| *ICE Facility* | 9 | 69% |
| *Correctional Facility* | 4 | 31% |

Percentages for types of detention are out of total in detention (n = 13).

### Online Training Module

Evaluators participated in optional surveys administered before (n = 23) and after (n = 18) the online training module described above. The questionnaires asked about prior experience, comfort with, and concerns about conducting forensic mental health evaluations by telephone (Supplemental Digital Appendix 1). Most respondents (n = 19, 83%) to the pre-module survey reported that they had never conducted forensic evaluations of asylum seekers by telephone. Participants were asked to rate their comfort with conducting remote forensic health evaluations of asylum seekers on a five-point Likert Scale (1 = “Not at all comfortable,” 5 = “Very comfortable”) before and after viewing the module. Of participants who completed both pre- and post-module surveys, eight (44%) respondents indicated an initial comfort level of four or five, with no change in comfort level between pre- and post-module surveys. Ten (56%) respondents who completed the pre-and post-module surveys indicated a comfort level of two or three on the pre-module survey; the comfort level reported by these ten respondents increased on average by 1.2 (0.4) points after viewing the module. Overall, most respondents (n = 16, 89%) to the post-module survey rated their comfort level a four or five. Of note, two respondents—who had received training but had never conducted any forensic medical evaluations—rated their comfort level a three in the post-module survey. Per the onboarding pathways described above, these two evaluators will shadow and work with a mentor evaluator in order to gain experience and confidence in their skills before performing remote evaluations independently.

### Next Steps

The initial goals of this pilot were to recruit and train personnel and to coordinate remote forensic mental health evaluations across several geographies. Next, we will formally evaluate the program. Although attorney feedback on work products has been positive, we plan to assess the quality of the affidavits resulting from our evaluations by performing a quantitative analysis comparing affidavits from participating clinicians’ remote and in-person evaluations. Further, we are tracking the legal outcomes of our cases and will analyze qualitative data and survey feedback from immigration attorneys about the impact of affidavits on these cases. Ultimately, future program evaluation must assess evaluees’ experiences with remote evaluations as an additional quality metric. International literature suggests that refugees and asylum seekers report high levels of satisfaction with tele-psychiatry, but further research is needed to confirm these findings in the specific population our program serves.^7,8^ Related research will also complement program evaluation by contributing to a limited evidence base about the efficacy of remote evaluations. Our program is conducting an ongoing qualitative study of the attitudes of legal professionals towards forensic mental health evaluations conducted by telephone. This study will provide greater insight into the utility and appropriateness of telephonic evaluations.

The initial experiences of the Remote Evaluation Network suggest that concerted coordination of forensic mental health evaluations by telephone or video improves access to forensic evaluations and provides a feasible alternative for asylum seekers unable to obtain in-person evaluations. If formal program evaluation affirms the quality of our pilot’s services and the acceptability of these evaluations to legal decision-makers, we will attempt to expand our program’s reach to additional low-resource geographies. Future research should survey the nationwide availability of pro bono forensic medical services to guide efficient, equitable allocation of remote services. While continuing to serve individuals detained in the U.S., we hope to facilitate evaluations for those who are forced, under the current federal Migrant Protection Protocols (MPP), to wait in Mexican border cities in unsafe conditions while their legal cases are pending.

The scope of the remote network is limited because MSHRP is currently only able to work with clients who have direct legal representation. By some estimates, just 14% of people in U.S. immigration detention have representation, and rates of representation for individuals who are barred from entering the U.S. under the MPP range from 0-11%.^9,10^ In the future, we hope to begin serving individuals participating in *pro se* (self-representation) legal clinics. Doing so would require careful consideration of practical and ethical questions as we seek to ensure access to and deployment of forensic evaluations for the most vulnerable individuals. For all individuals seeking protected immigration status—those in the community, detention centers, and Mexican border cities—we hope to build resources to facilitate local continuity care referrals addressing the mental health needs identified during forensic evaluations and to continue advocating for individuals to receive appropriate medical resources, especially in detention facilities.

## Data Availability

Data reported in this manuscript are not publicly available.

## Acknowledgments

The authors wish to thank Dr. Mary Rojas, the Medical Student Research Office, and Dean David Muller at the Icahn School of Medicine at Mount Sinai for their continuing support; Dr. Kim Baranowski for her leadership and guidance in building this program; Erik Popil, Dr. Gale Justin, and the Instructional Technology Group at the Icahn School of Medicine at Mount Sinai for their assistance in creating the Remote Evaluation Network’s online training module; and the MCJ Amelior Foundation for its generosity. The authors also wish to thank the clinicians in the Remote Evaluation Network and the pro bono immigration attorneys we have partnered with for their incredible commitment and dedication to making this pilot possible.

## Disclosures

### Ethical approval

The collection of anonymous survey data was determined to be exempt human research by the Mount Sinai Institutional Review Board.

### Disclaimers

None.

### Previous presentations

A description of the Remote Evaluation Network has been accepted for oral presentation at the North American Refugee Health Conference, Cleveland, Ohio, August 16, 2020.

## Tables and Supplemental Digital Appendix

## Supplemental Digital Appendix 1: Pre- and Post-Module Surveys

### Pre-Module Survey

Please select “I agree” and you will be taken to the survey. If you would like to skip the survey and proceed right to the module, please press the “Go to Video” button on the right hand side of the screen.

∘ I agree

Certification/degrees earned:

∘ LCSW
∘ MD Psychiatrist
∘ MD Other Specialty
∘ NP
∘ PhD Psychologist
∘ PsyD
∘ Other:____

Are you currently involved with, or in the process of onboarding with, Mount Sinai Human Rights Program’s Remote Evaluation Network? Please note that if you are not a current or prospective MSHRP evaluator, you are still welcome to participate in this survey.

∘ Yes
∘ No

How many years have you been in practice?

____

### Background on Asylum Work

Estimated number of mental health asylum evaluations I have conducted:

∘ 0
∘ 1-5
∘ 5-10
∘ 10-15
∘ 15+

Estimated number of mental health asylum evaluations I have conducted remotely (via telephone, video conference, or other platform):

∘ 0
∘ 1-5
∘ 5-10
∘ 10-15
∘ 15+

What type of training did you receive in conducting asylum evaluations?

____

### Attitudes towards tele-psychiatric evaluations

How comfortable are you conducting remote, telephonic or video psychiatric interviews?

> Not at all comfortable – O 1 O 2 O 3 O 4 O 5 – Very comfortable

How comfortable are you conducting remote, telephonic or video forensic psychiatric asylum evaluations?

> Not at all comfortable – O 1 O 2 O 3 O 4 O 5 – Very comfortable

Which of the following, if any, is a concern of yours in conducting remote asylum evaluations (select all that apply):

∘ Building rapport with patient
∘ Communicating via interpreter
∘ Capturing adequate clinical information
∘ Assessing elements of Mental Status Exam (i.e. affect, appearance, motor)
∘ Assessing credibility of client
∘ Managing patient suicidality
∘ Legal considerations (i.e. license not applying in other states)
∘ Other: ____

Please choose a 5-digit identification code, which will enable us to link your responses in the pre-module survey with your responses in the post-module survey. This allows us to connect your responses, while keeping your identity anonymous. We recommend you use something easy to remember, as we will ask for it in the post-module survey. Please avoid using a generic code (e.g. “12345”).

____

### Post-Module Survey

Please input the 5-digit identification code you chose during the pre-module survey.

### Attitudes towards tele-psychiatric evaluations

Please rate the degree to which you agree with the following: This module was helpful in preparing me for telephonic forensic asylum evaluations.

> Not at all helpful – O 1 O 2 O 3 O 4 O 5 – Very helpful

Please rate your comfort conducting forensic psychiatric asylum assessments remotely (via telephone, video conference, or other platform).

> Not at all comfortable – O 1 O 2 O 3 O 4 O 5 – Very comfortable

How likely are you to apply what you have learned in your future forensic asylum evaluations?

> Not at all likely – O 1 O 2 O 3 O 4 O 5 – Very likely

If you felt that “Building rapport with patient” was a concern, please rate the extent to which this module addressed this concern:

> Did not effectively address this concern – O 1 O 2 O 3 O 4 O 5 – Very effectively addressed concern.

If you felt that “Communicating via interpreter” was a concern, please rate the extent to which this module addressed this concern:

> Did not effectively address this concern – O 1 O 2 O 3 O 4 O 5 – Very effectively addressed concern.

If you felt that “Capturing adequate clinical information” was a concern, please rate the extent to which this module addressed this concern:

> Did not effectively address this concern – O 1 O 2 O 3 O 4 O 5 – Very effectively addressed concern.

If you felt that “Assessing elements of Mental Status Exam” was a concern, please rate the extent to which this module addressed this concern:

> Did not effectively address this concern – O 1 O 2 O 3 O 4 O 5 – Very effectively addressed concern.

If you felt that “Assessing credibility of client” was a concern, please rate the extent to which this module addressed this concern:

> Did not effectively address this concern – O 1 O 2 O 3 O 4 O 5 – Very effectively addressed concern.

Was there anything you would like to have seen addressed in this module that was not covered?

____

